# Biomechanical outcomes of the pendulum test characterize individual differences in activated versus resting leg rigidity in people with Parkinson’s disease

**DOI:** 10.1101/2020.09.07.20188474

**Authors:** Giovanni Martino, J. Lucas McKay, Stewart A. Factor, Lena H. Ting

## Abstract

Leg rigidity is associated with frequent falls in people with Parkinson’s disease (PD), suggesting a potential role in functional balance and gait impairments. Changes in neural state due to secondary tasks, e.g. activation maneuvers, can exacerbate (or “activate”) rigidity, possibly increasing the risk of falls. However, the subjective interpretation and coarse classification of the standard clinical rigidity scale has prohibited the systematic, objective assessment of resting and activated leg rigidity. The pendulum test is an objective diagnostic method that we hypothesized would be sensitive enough to characterize resting and activated leg rigidity.

We recorded kinematic data during the pendulum test in 15 individuals with PD, spanning a range of leg rigidity severity (slight to severe). From the recorded data of leg swing kinematics we measured biomechanical outcomes including first swing excursion, first extension peak, number and duration of the oscillations, resting angle, relaxation index, maximum and minimum angular velocity. We examined associations between biomechanical outcomes and clinical leg rigidity score. We evaluated the effect of increasing rigidity through activation maneuvers on biomechanical outcomes. Finally, we assessed whether either biomechanical outcomes or changes in outcomes with activation were associated with fall history.

Our results suggest that the biomechanical assessment of the pendulum test can objectively quantify leg rigidity among people with PD. We found that the presence of marked rigidity during clinical exam significantly impacted biomechanical outcomes, i.e. first extension peak, number of oscillations, relaxation index and maximum angular velocity. No differences in the effect of activation maneuvers between groups with clinically assessed moderate and marked rigidity were observed, suggesting that activated rigidity may be independent of resting rigidity and should be scored as independent variables. Moreover, we found that fall history was more common among people whose rigidity was increased with a secondary task, as measured by biomechanical outcomes.

We conclude that different mechanisms contributing to resting and activated rigidity may play an important yet unexplored functional role in balance impairments. The pendulum test may contribute to a better understanding of fundamental mechanisms underlying motor symptoms in PD, evaluating the efficacy of treatments, and predicting the risk of falls.

## 1 Introduction

Rigidity is a cardinal feature of Parkinson’s disease (PD) and its role in functional balance and gait impairment has been questioned (Wright et al., 2007; Franzén et al., 2009). Our recent work suggested that leg–but not arm, neck, or total–rigidity score is associated with frequent falls in people with PD (McKay et al., 2019). However, leg rigidity scores reflect a coarse and subjective categorization based on subitem 3.3 in the Movement Disorders Society Unified Parkinson’s Disease Rating Scale (MDS-UPDRS). Rigidity is clinically described as a constant increased resistance to a passive or externally induced motion throughout the range of movement (Fung and Thompson, 2002). Rigidity generally responds well to dopaminergic medication and surgical interventions (Xia, 2011), therefore representing an indicator of treatment response in clinical management and may also be used as a metric for pain and impaired mobility, as patients perceive rigidity as aching and stiffness in the muscles and joints.

Changes in neural state can exacerbate rigidity, but such effects are quantified only at the lowest range of the MDS-UPDRS. In the MDS-UPDRS, a passive movement is imposed by an examiner and the perceived stiffness rated with an ordinal score from 0 (absent rigidity) to 4 (severe rigidity) for each arm, leg, and for the neck. In the ‘resting rigidity’ condition, the subject is asked to completely relax during the assessment (Webster and Mortimer, 1977). An activation maneuver (such as finger tapping) is used in the MDS-UPDRS to evaluate ‘activated rigidity’ only if a person exhibits no resistance when relaxed; thus activation maneuver are mainly used in only at the mildest rigidity levels (Fung et al., 2000; Powell et al., 2011). Moreover, activated rigidity has not been systematically studied in the leg, although it could may play a causal role in falls (McKay et al., 2019). Thus, more sensitive and objective methods for quantifying leg rigidity are necessary to enable associations between rigidity and other biomechanical or clinical outcomes.

Here, we hypothesized that the pendulum test would be an objective, sensitive, and practical test to characterize resting and activated rigidity based on its biomechanical outcomes and electromyographic (EMG) recordings. Various methods have been proposed in literature to objectively quantify rigidity in PD (Eisen, 1987; Andreeva and Khutorskaya, 1996; Kirollos et al., 1996; Patrick et al., 2001; Marusiak et al., 2010; Xia et al., 2011; Powell et al., 2012; Endo et al., 2015; Zetterberg et al., 2015), but focus has been on the upper limbs, and they have not been implemented in the clinical setting because of their complexity, need for expensive devices, and time involved. In contrast, the pendulum test is a diagnostic method that allows passive joint resistance to be objectively characterized based on the the pattern of lower leg movement after release from the horizontal (Wartenberg, 1951). Assessment using the pendulum test has been shown to be sensitive to standard clinical measurements of spasticity in children with cerebral palsy (Fowler et al., 2000; Jr and Miller, 2004; Szopa et al., 2014), multiple sclerosis patients (Bianchi et al., 1999), and stroke survivors (Brown et al., 1988; Lin and Rymer, 1991; Bohannon et al., 2009), in which the decrease in the first swing excursion, number of oscillations and resting angle, along with abnormal burst of activation in quadriceps and hamstrings has been found to be the best predictor of spasticity severity. Furthermore, the use of a computational model associated with pendulum test data has been shown to be capable of dissociating the contributions of abnormal muscle tone versus abnormal reflex excitability to spasticity (De Groote et al., 2018), revealing new insights into physiological mechanisms of spasticity. In De Groote et al. we suggested that the abnormal limb motion in children with cerebral palsy results from the interactions between muscle tone and the resulting short-range stiffness, and force-dependent reflexes. In PD, marked reductions in leg swing velocity and resting angle have been observed (Brown et al., 1988) and attributed to increased damping in simulations (Le Cavorzin et al., 2003). However, these reductions have not been associated with the degree of leg rigidity.

The pendulum test may also be sufficiently sensitive to test the level of activated rigidity which we hypothesized could potentially increase the risk of falling during activities of daily living (ADL’s). Several studies have shown that the presence of a secondary task or activation maneuver considerably enhances rigidity (Kelly et al., 2012). The degree of the increase in rigidity with activation can differ from patient to patient and can be present in both on- and off- dopaminergic medication states (Fung et al., 2000; Hong et al., 2007; Shapiro et al., 2007; Powell et al., 2011). Also, different medications and dosages have been reported to have variable effects on both resting and activated rigidity (Webster and Mortimer, 1977; Kirollos et al., 1996; Relja et al., 1996; Krack et al., 2003; Shapiro et al., 2007), suggesting that different neural mechanisms could play a role in the manifestation of rigidity. However, the difference between activated and resting rigidity and its relationship with the degree of severity of rigidity at rest or to other clinical outcomes has not been explored before.

We hypothesized that both resting and activated rigidity in PD alter pendulum test kinematics and EMG patterns. We predicted that the biomechanical outcomes of the pendulum test, namely first swing excursion, first extension peak, number and duration of the oscillations, resting angle,relaxation index, maximum and minimum angular velocity, would be indicators of leg rigidity severity in people with PD. We further predicted that an activation maneuver would alter pendulum test outcomes, but that the effects would vary from an individual to the next. Finally we tested whether the level of activated rigidity would could be associated with fall history.

## 2 Materials and Methods

### 2.1 Study participants

We performed the pendulum test in fifteen participants with PD. Participants were recruited from the cohort of an observational 1-year fall risk study (McKay et al., 2019). We included patients with a diagnosis of clinically defined PD who exhibited rigidity during MDS-UPDRS-III testing in the practically-defined “OFF” state (see below). Exclusion criteria were history of musculoskeletal and/or neurological disorders other than PD, inability to walk ≥3 meters with or without assistance and advanced stage dementia in which patients were unable to perform activities of daily living independently, signs of spasticity or paratonia at clinical examination. Sample size was selected to meet or exceed common recommendations of ≈10 cases/independent variable in regression analyses (Vittinghoff and McCulloch, 2007) and ≥12 cases/group in preclinical studies (Julious, 2005). PD participants were assessed in the practically defined OFF medication state, ≥12 hours after their last dose of antiparkinsonian medications (Langston et al., 1992). Each participant’s neurologist signed an OFF-medication clearance form before the patient was asked to withhold their medications for the purpose of this experiment. All participants provided written informed consent prior to participation according to protocols approved by the Institutional Review Board of Emory University.

Lower limb rigidity was evaluated at the beginning of the experimental session by a trained examiner, following the MDS-UPDRS guidelines: rigidity in the lower extremities was tested by fully extending and flexing the knee with the patient sitting (0 = Absent, 1 = Slight or detectable only when activated by mirror or other movements, 2 = Mild to moderate, 3 = Marked, but full range of motion easily achieved, 4 = Severe, range of motion achieved with difficulty). The participants were classified as “fallers” if they reported cases of falls in the six month period before the data collection and were classified as “non-fallers” otherwise (McKay et al., 2019).

### 2.2 Pendulum test

The pendulum test was performed with the subject sitting on a treatment table with the trunk inclined approximated 40° from the vertical to provide a comfortable starting position (Stillman and McMeeken, 1995). We designed a custom backrest that fits on a physical therapy table to control the posture of the participants. During the test, the examiner dropped the lower leg of the participant from the horizontal position with an extended knee joint; the lower leg was then allowed to swing freely under the influence of gravity. In each participant the pendulum test was assessed during four randomized different conditions: a baseline condition, with the subject completely relaxed and with the hands on his/her lap, and while performing three different activation maneuvers (described below). The most rigid lower limb was assessed for each participant. Three trials were performed for each condition and a pause of 40s was ensured between them to avoid fatigue due to the activation maneuver. The same examiner carried out the test across all the sessions and participants.

### 2.3 Activation maneuvers

We first identified which activation maneuver was most effective in increasing rigidity during the pendulum test. We tested the effect of three different activation maneuvers: finger tapping, fist clenching, and the Jendrassik maneuver. The rationale for the incorporation of an activation maneuver lies in that activation maneuver has been shown to enhance the degree of rigidity in PD patients (Matsumoto et al., 1963; Kelly et al., 2012). The finger tapping test is one of the standard activation maneuvers used to clinically evaluate rigidity in PD (Shimoyama et al., 1990; Martínez-Martín et al., 1994) and is indicated as one of the activation maneuvers used to assess rigidity in the UPDRS scale (Fahn and Elton, 1987). The second activation maneuver consists in a sustained clenching of the fists (Meara and Cody, 1992). As an alternative to finger tapping and clenching, the Jendrassik maneuver is a common clinical test where the patient interlocks the fingers of each hand in hook-like fashion and isometrically pulls the hands apart as strongly as possible (Ertuglu et al., 2018).

### 2.4 Data analysis

Joint kinematics were recorded by means of a motion capture analysis system (Vicon). Participants wore a 25-marker set according to a modified version of the Vicon’s Plug-in Gait model (Welch and Ting, 2008). Kinematic data were filtered using a 2^nd^ order zero lag low pass Butterworth with a cut off frequency of 5Hz. Knee angles were taken by measuring the absolute angle of the leg segment in the sagittal plane. Biomechanical outcomes (Fig. 1B) were then calculated including: first swing excursion (FSE), first extension peak (FPE), number (N) and duration (d) of the oscillations, resting angle (θ_rest_), relaxation index (RI), maximum (V_max_) and minimum (V_min_) angular velocity. The end of the oscillation was calculated by considering a cutoff of 3° toward extension (Fowler et al., 2000). We also recorded EMG activity from biceps femoris (BF) and rectus femoris (RF) in a subset of participants. EMG data were high-pass filtered (35Hz, 3rd order zero lag Butterworth filter), demeaned, rectified and low-pass filtered (40Hz).

**Figure 1.**
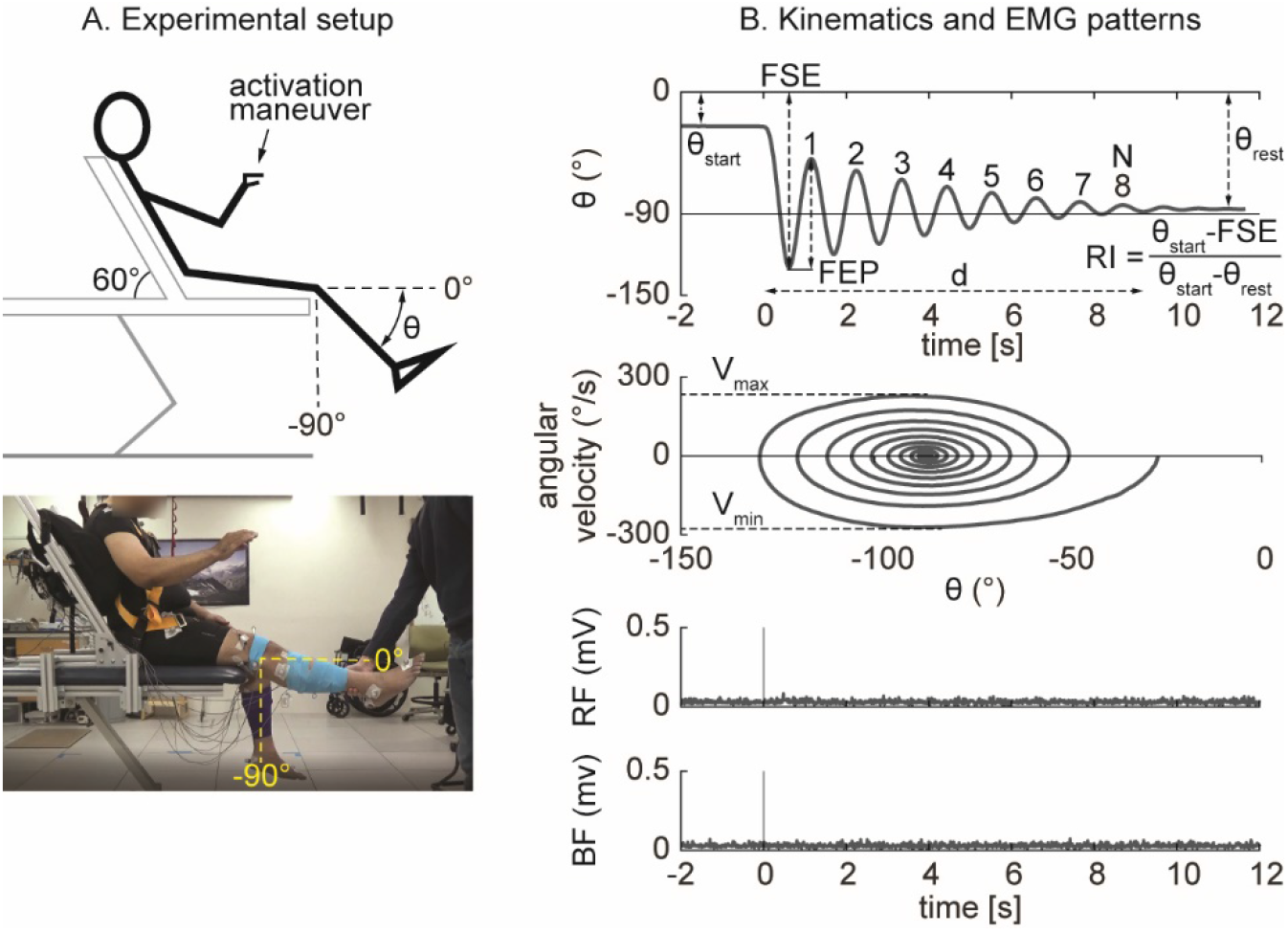
Experimental setup and outcomes of the pendulum test. The pendulum test was performed with the subject sitting on a treatment table with the trunk inclined approximated 60° from the horizontal to provide a comfortable starting position (**A**). The swinging leg behaves as a damped pendulum, oscillating several times before coming to rest. First swing excursion (FSE), number (N) and duration (d) of the oscillations, first extension peak (FEP), resting angle (θ_rest_), maximum (V_max_) and minimum (V_min_) angular velocity were assessed from kinematic data (**B**). The middle panels show the typical ‘whirlpool’ pattern of angular velocity against angle data. EMG activity of rectus femoris (RF) and biceps femoris (BF) were also recorded in a subset of participants (bottom panel).

**Figure 2.**
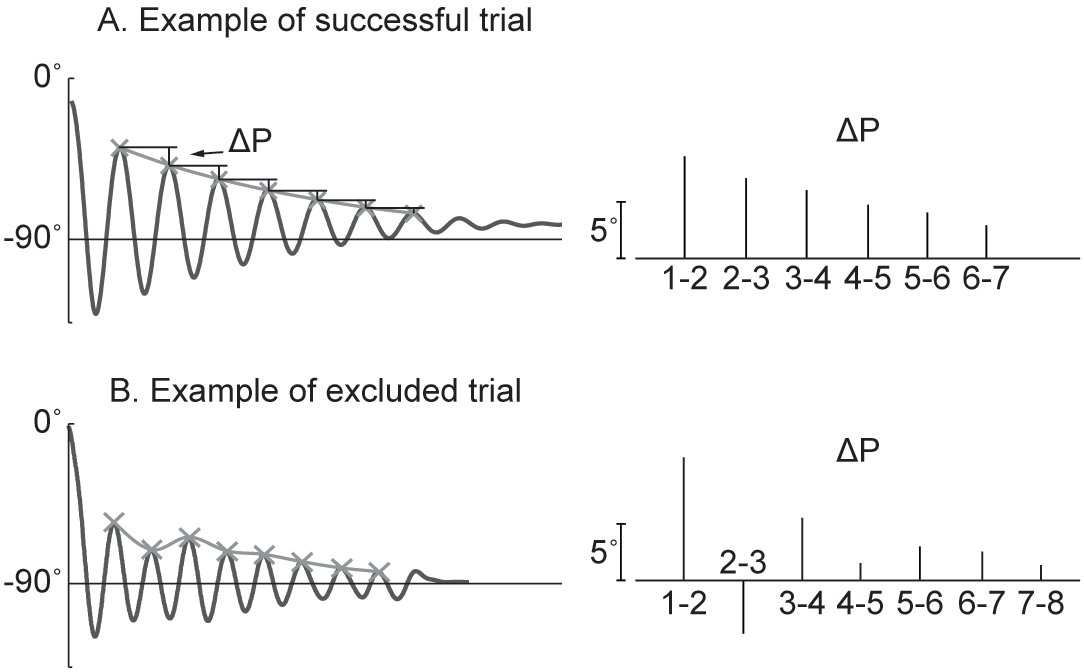
Example of successful and excluded trials. The pattern of the knee angle during the pendulum test follows an exponential decrease of the peaks (**A**). When peaks variation increase it indicates voluntary input of the participant (**B**). We excluded the trials in which the decrement from the i-th peak to i-th+1 is not greater than the following one.

### 2.5 Statistical analysis

Participants were classified as either slight-to-moderate rigidity (leg rigidity score from 1 to 2) or marked-to-severe rigidity (leg rigidity score from 3 to 4). Between-groups differences in clinical and demographic variables were assessed with t-tests and Chi-squared tests as appropriate. Between-groups differences in outcome measures were assessed with independent samples *t*-tests. Within-subject differences in outcome measures between the activated and resting states were assessed with paired-samples *t*-tests. Differences in clinical and demographic variables between fallers and non-fallers were assessed with t-tests and Chi-squared tests as appropriate. P-values <0.05 were considered statistically significant. Due to the exploratory nature of the study no corrections for multiple comparisons were used.

## 3 Result

Fifteen participants with PD (11 males and 4 females, mean age 67±10 years) enrolled in the study. Demographic and clinical characteristics are shown in Table 1. No significant differences in clinical or demographic characteristics were observed between the low and high rigidity groups (Table 2). Consistent with previous report (McKay et al., 2018), some significant differences were observed between fallers and non-fallers on Sex, Total MDS-UPDRS-III score, rigidity score, and LED (Table 3). We excluded from the analysis all the trials in which the participants were unable to relax the leg during the test. Three subjects were unable to relax during the whole session and were excluded from further analysis. The number (mean±SD) of successful trials among participants was 2±1 during resting state, 1±1 during finger tapping, 2±1 during fist clenching, and 2±1 during the Jendrassik maneuver. Initial analyses (one-way ANOVA, post-hoc Tukey-Kramer) identified no significant differences between the effects of the three different activation maneuvers on the biomechanical outcomes (all p > 0.05). Therefore we aggregated the results of all of the activated conditions.

**Table 1.**
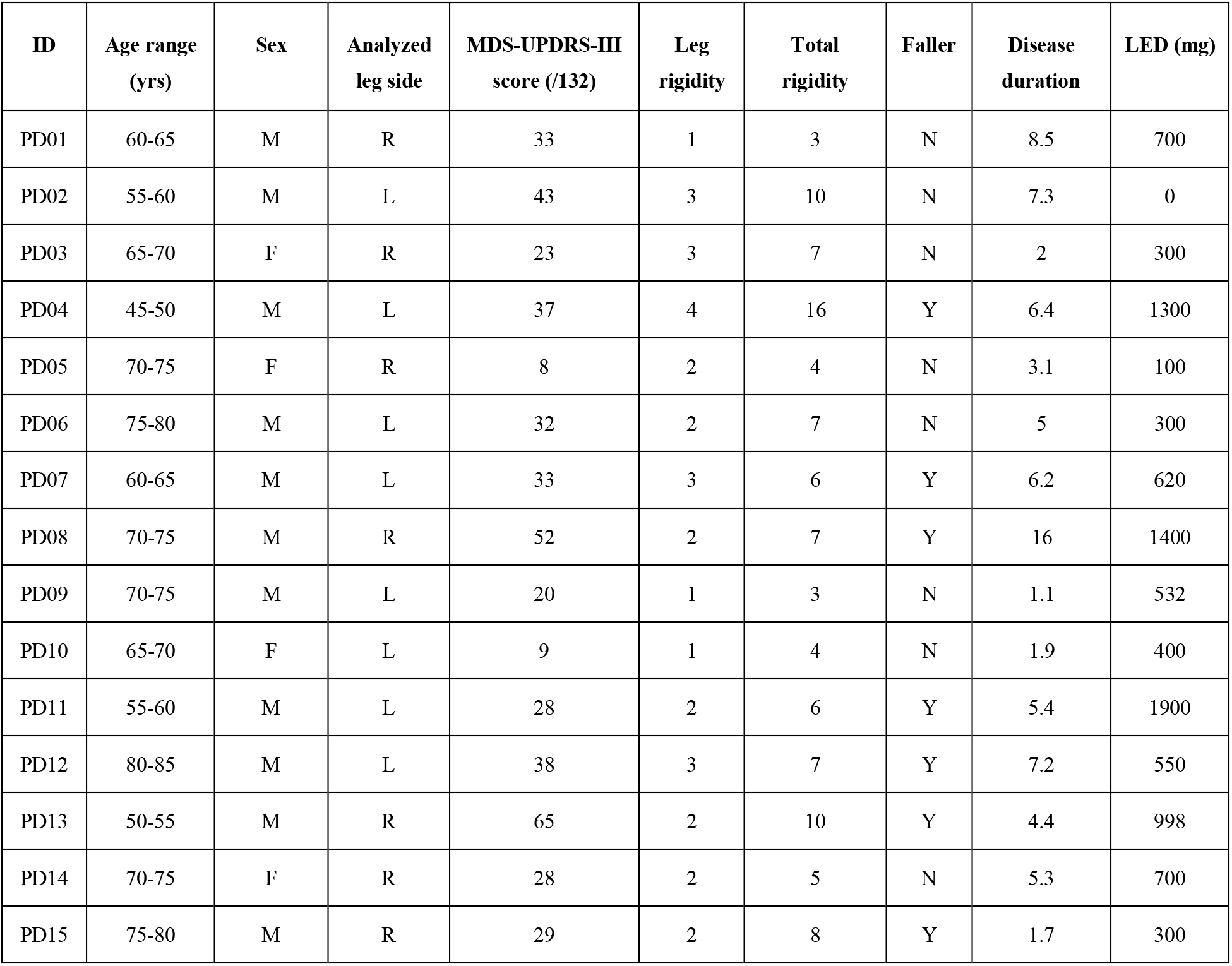
Demographic and clinical characteristics of the study participants.

| **ID** | **Age range**<br>**(yrs)** | **Sex** | **Analyzed**<br>**leg side** | **MDS-UPDRS-III**<br>**score (/132)** | **Leg**<br>**rigidity** | **Total**<br>**rigidity** | **Faller** | **Disease**<br>**duration** | **LED (mg)** |
| --- | --- | --- | --- | --- | --- | --- | --- | --- | --- |
| PD01 | 60-65 | M | R | 33 | 1 | 3 | N | 8.5 | 700 |
| PD02 | 55-60 | M | L | 43 | 3 | 10 | N | 7.3 | 0 |
| PD03 | 65-70 | F | R | 23 | 3 | 7 | N | 2 | 300 |
| PD04 | 45-50 | M | L | 37 | 4 | 16 | Y | 6.4 | 1300 |
| PD05 | 70-75 | F | R | 8 | 2 | 4 | N | 3.1 | 100 |
| PD06 | 75-80 | M | L | 32 | 2 | 7 | N | 5 | 300 |
| PD07 | 60-65 | M | L | 33 | 3 | 6 | Y | 6.2 | 620 |
| PD08 | 70-75 | M | R | 52 | 2 | 7 | Y | 16 | 1400 |
| PD09 | 70-75 | M | L | 20 | 1 | 3 | N | 1.1 | 532 |
| PD10 | 65-70 | F | L | 9 | 1 | 4 | N | 1.9 | 400 |
| PD11 | 55-60 | M | L | 28 | 2 | 6 | Y | 5.4 | 1900 |
| PD12 | 80-85 | M | L | 38 | 3 | 7 | Y | 7.2 | 550 |
| PD13 | 50-55 | M | R | 65 | 2 | 10 | Y | 4.4 | 998 |
| PD14 | 70-75 | F | R | 28 | 2 | 5 | N | 5.3 | 700 |
| PD15 | 75-80 | M | R | 29 | 2 | 8 | Y | 1.7 | 300 |

**Table 2.** Demographic and clinical characteristics of the study participants, overall and stratified on rigidity status.

|  | Low Rigidity (N=10) | High Rigidity (N=5) | Entire Sample (N=15) | P value |
| --- | --- | --- | --- | --- |
| Age |  |  |  | 0.442 |
| Mean (SD) | 68 (9) | 63 (12) | 66 (10) |  |
| Range | 51 - 80 | 47 - 81 | 47 - 81 |  |
| Sex |  |  |  | 0.68 |
| F | 3 (30.0%) | 1 (20.0%) | 4 (26.7%) |  |
| M | 7 (70.0%) | 4 (80.0%) | 11 (73.3%) |  |
| MDS-UPDRS-III |  |  |  | 0.605 |
| Mean (SD) | 30 (17) | 35 (7) | 32 (15) |  |
| Range | 8 - 65 | 23 - 43 | 8 - 65 |  |
| Total Rigidity† |  |  |  | 0.189 |
| Mean (SD) | 4.0 (2.0) | 6.0 (3.7) | 4.7 (2.7) |  |
| Range | 2 - 8 | 3 - 12 | 2 - 12 |  |
| Faller |  |  |  | 0.464 |
| N | 6 (60.0%) | 2 (40.0%) | 8 (53.3%) |  |
| Y | 4 (40.0%) | 3 (60.0%) | 7 (46.7%) |  |
| PD Duration |  |  |  | 0.787 |
| Mean (SD) | 5.2 (4.8) | 5.8 (2.2) | 5.4 (3.7) |  |
| Range | 1.1 - 16.0 | 2.0 - 7.3 | 1.1 - 16.0 |  |
| LED |  |  |  | 0.553 |
| Mean (SD) | 733 (558) | 554 (482) | 673 (524) |  |
| Range | 100 - 1900 | 0 - 1300 | 0 - 1900 |  |

†Total rigidity omits leg rigidity score.

**Table 3.**
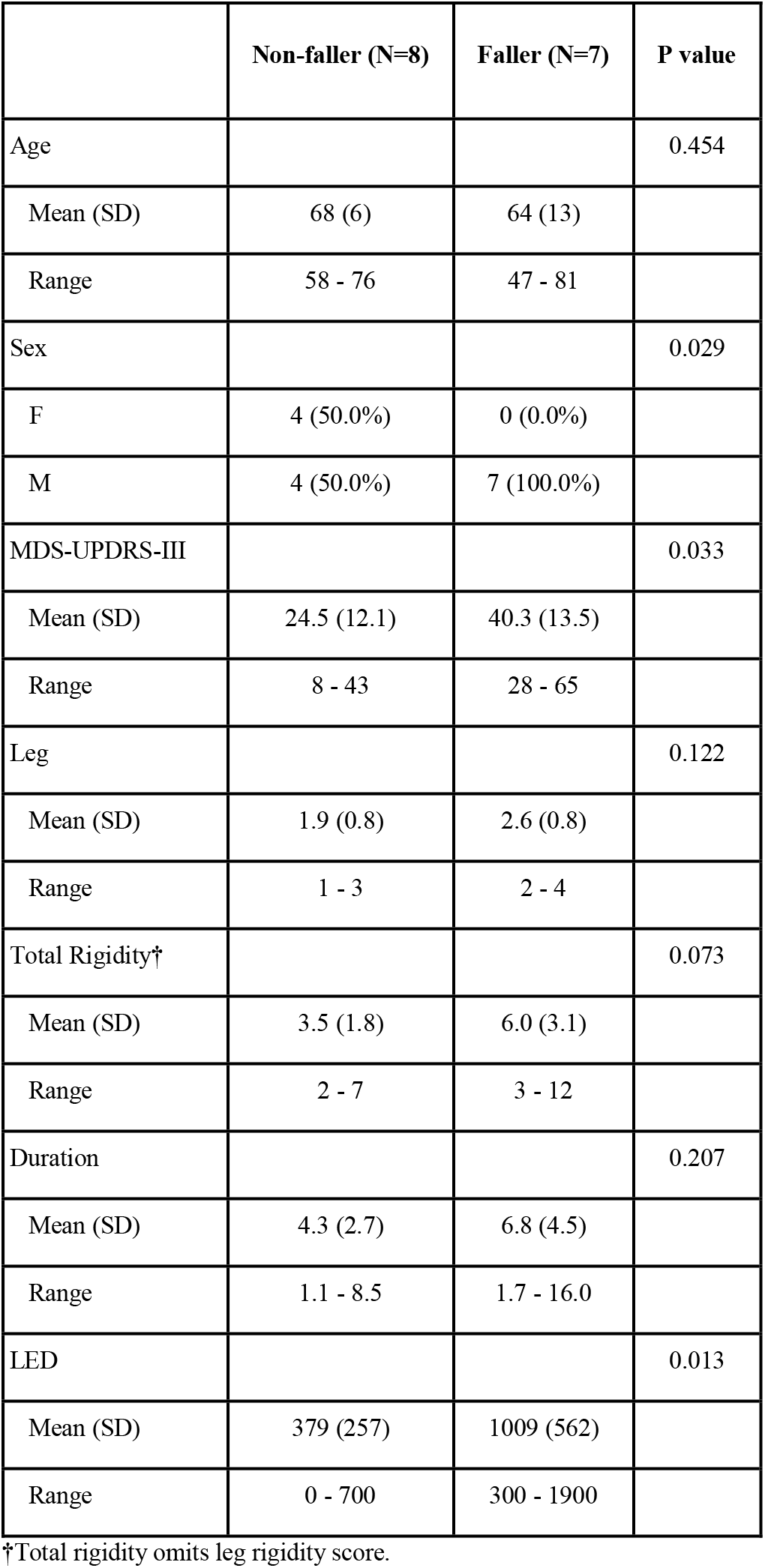
Demographic and clinical characteristics of the study participants stratified on prevalence of previous falls.

|  | Non-faller (N=8) | Faller (N=7) | P value |
| --- | --- | --- | --- |
| Age |  |  | 0.454 |
| Mean (SD) | 68 (6) | 64 (13) |  |
| Range | 58 - 76 | 47 - 81 |  |
| Sex |  |  | 0.029 |
| F | 4 (50.0%) | 0 (0.0%) |  |
| M | 4 (50.0%) | 7 (100.0%) |  |
| MDS-UPDRS-III |  |  | 0.033 |
| Mean (SD) | 24.5 (12.1) | 40.3 (13.5) |  |
| Range | 8 - 43 | 28 - 65 |  |
| Leg |  |  | 0.122 |
| Mean (SD) | 1.9 (0.8) | 2.6 (0.8) |  |
| Range | 1 - 3 | 2 - 4 |  |
| Total Rigidity† |  |  | 0.073 |
| Mean (SD) | 3.5 (1.8) | 6.0 (3.1) |  |
| Range | 2 - 7 | 3 - 12 |  |
| Duration |  |  | 0.207 |
| Mean (SD) | 4.3 (2.7) | 6.8 (4.5) |  |
| Range | 1.1 - 8.5 | 1.7 - 16.0 |  |
| LED |  |  | 0.013 |
| Mean (SD) | 379 (257) | 1009 (562) |  |
| Range | 0 - 700 | 300 - 1900 |  |

†Total rigidity omits leg rigidity score.

### 3.1 Examples of pendulum test kinematic patterns

Different kinematic patterns of the pendulum test were observed across lower leg rigidity scores. For example, in a participant with slight leg rigidity (Fig. 3A, score = 1/4) the leg oscillated four times, with a first swing excursion of greater than 100°, and negative peak angular speed of about –300°/s. A participant with mild to moderate rigidity (score = 2/4) exhibited a similar pattern (Fig. 3B), with the leg oscillating five times before coming to rest. Although a participant with marked rigidity (Fig. 3C, score = 3/4) also had about four leg oscillations, the first swing excursion was smaller than participants with lower rigidity scores, near 90°, and negative peak angular speed of about –230°/s. In the participant with severe rigidity (Fig. 3D, score = 4/4) no oscillations were observed, with the leg slowly lowering to a less vertical resting angle that other participants.

**Figure 3.**
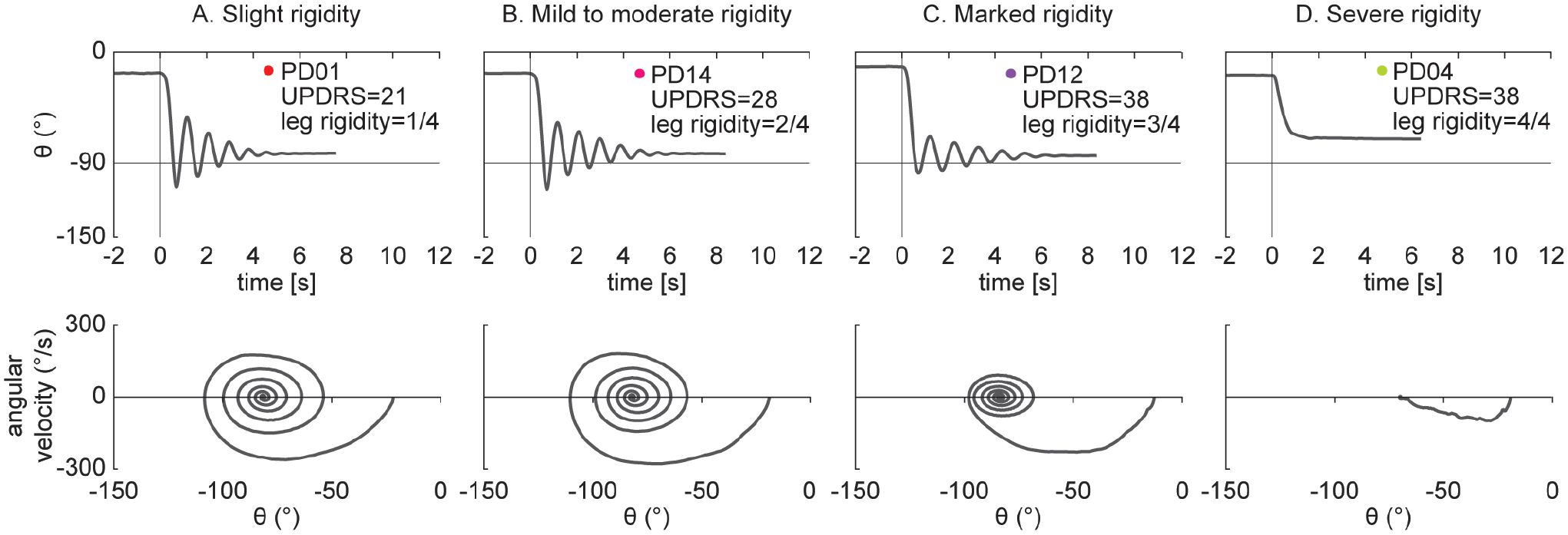
Example of pendulum test kinematic traces in four PD individuals with increasing levels of lower leg rigidity (as measured by following the UPDRS guidelines). Slight rigidity (**A**). Mild to moderate rigidity (**B**). Marked rigidity (**C**). Severe rigidity (**D**).

### 3.2 Low vs high rigidity scores

We found differences in the biomechanical outcomes of the pendulum test between PD participants with slight to moderate leg rigidity scores (1 to 2) and with marked to severe leg rigidity scores (3 to 4) during resting condition (Fig. 4). As rigidity increased there was a significant reduction of the first extension peak (Fig. 4B, 58°±15 vs 34°±23, p = 0.042), number of oscillations (Fig. 4C, 5±1 vs 3±2, p = 0.047), relaxation index (Fig. 4E, 1.5±0.1 vs 1.3±0.2, p = 0.013) and maximum angular velocity (Fig. 4G, 182°/s±35 vs 105°/s±69, p = 0.019). Furthermore, most of the individual values for both groups fell out of the range of the mean (±SD) of the biomechanical parameters (Fig. 4, grey areas) estimated from previously reported pendulum test data in healthy subjects (Stillman and McMeeken, 1995).

**Figure 4.**
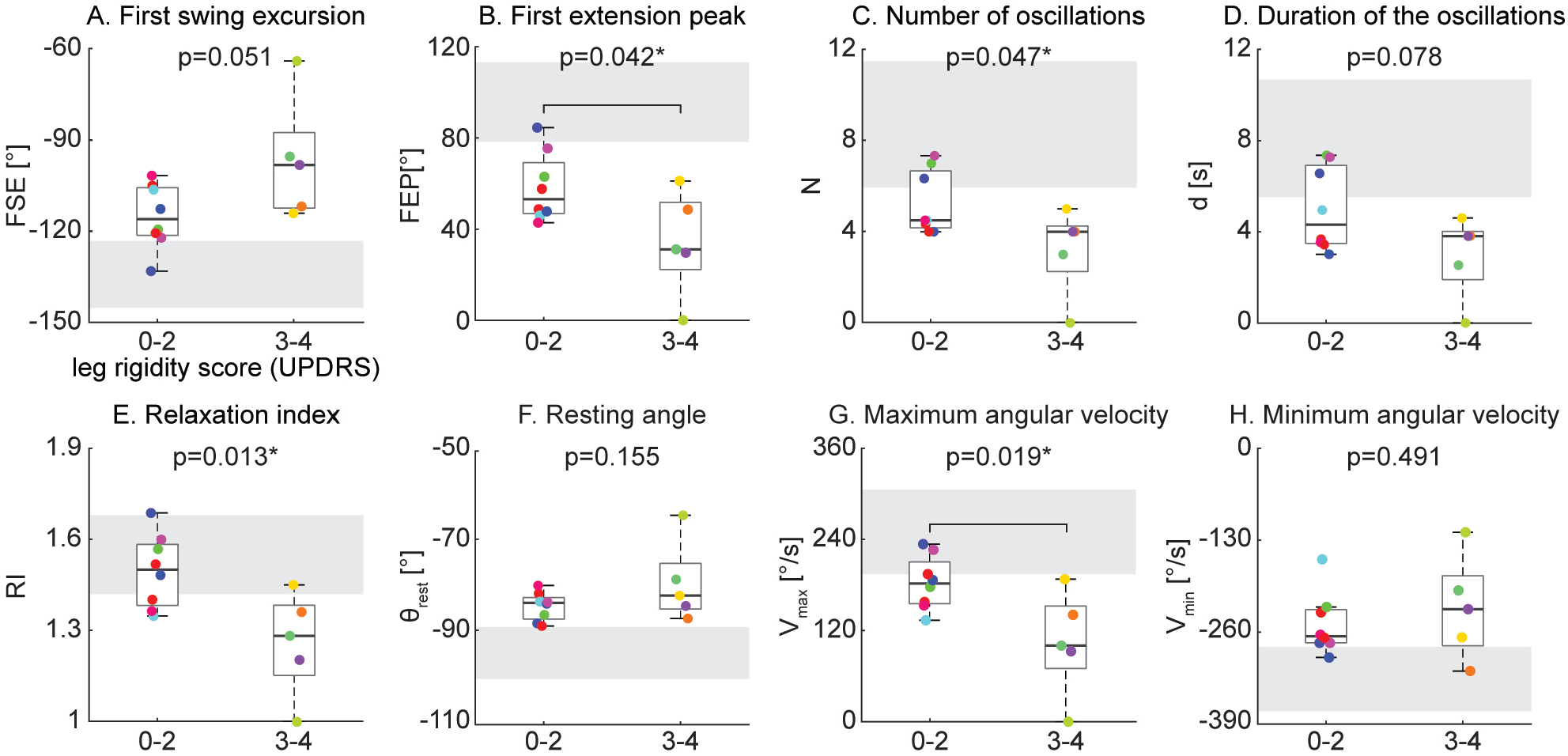
Kinematic outcomes of the pendulum test in the baseline condition. Subjects were grouped based on rigidity score of the recorded leg: subjects with leg rigidity score from 0 to 2 (absent to moderate rigidity) and subjects with leg rigidity score from 3 to 4 (marked to severe rigidity). Grey areas correspond to mean ±SD of biomechanical outcomes for healthy subjects estimated from Stillman and McMeeken (1995). Asterisks denote significant values (p< 0.05).

### 3.3 Effects of activation maneuver

Individual differences in the effects of the activation maneuver were observed, even across participants with similar rigidity scores. For example, three individuals with the same rigidity score exhibited marked differences in whether and how biomechanical outcomes changed in the presence of an activation maneuver (Fig. 5A-C, score = 2/2). Participant A (Fig. 5A) exhibited 8 oscillations of the leg during the resting condition and no changes in the kinematics during the activated condition, though increased BF tonic activity was observed prior to the movement. Although the other two participants with a leg rigidity score of 2 (Fig. 5B-C) had a similar number of oscillations in the resting condition (N = 4) that was reduced during an activation maneuver (N = 3), they exhibited differences in other features of the pendulum test outcomes. During an activation maneuver in participant B (Fig. 5B) first extension peak and maximum velocity decreased, tonic activity in both the RF and BF muscles increased, and reflexive activity in the BF was observed during the first knee extension. In Participant C (Fig. 5C) a decrease in the first swing excursion, resting angle and minimum and maximum angular velocity was observed during an activation maneuver, together with increased tonic activity in RF. Participant D had severe rigidity (Fig. 5D) and did not exhibit any oscillations in either the resting or activated states, but angular velocity decreased in the presence of an activation maneuver. We also observed a change in resting angle after the end of the activation maneuver in the most severe subject (Fig. 5D).

**Figure 5.**
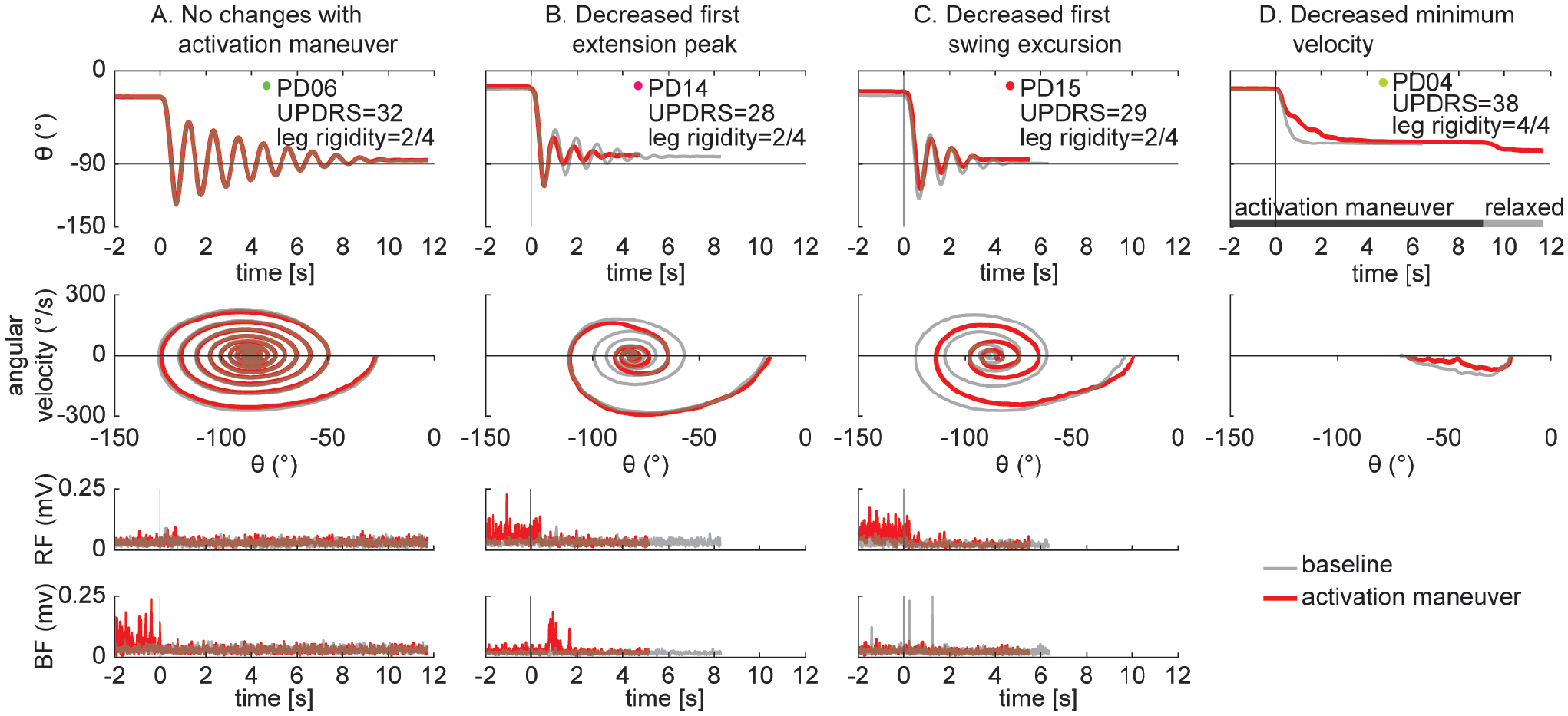
Individual specific changes in the pattern of leg movement and EMG activity among PD subjects while performing an activation maneuver (AM). In subject PD06 we found no kinematic changes with an activation maneuver (**A**). In subject PD14 we found a decrease of the first extension peak and of the number and duration of the oscillations during AM (**B**). In subject PD15 we found a decrease in the first swing excursion and of the number and duration of the oscillations during AM (**C**). In the subject with severe rigidity (PD04) we found a decrease of the angular velocity of the leg during AM. No EMG recorded (**D**).

The changes in three biomechanical outcomes in the activation versus resting state were found having a distribution with mean significantly different from zero, but the magnitude of this effect did not depend on the severity of leg rigidity (Fig. 6). A one-sample t-test revealed a significant effect of activation on the first extension peak (Fig. 6B, p = 0.018), number of oscillations (Fig. 6C, p = 0.013) and duration of the oscillations (Fig. 6D, p = 0.013). A two-sample t-test did not reveal any significant difference in the effect of activation maneuver on the biomechanical outcomes of the pendulum test when comparing the group with absent to moderate rigidity (0–2) to the group with marked to severe rigidity (3–4), (Fig. 6, all p > 0.05).

**Figure 6.**
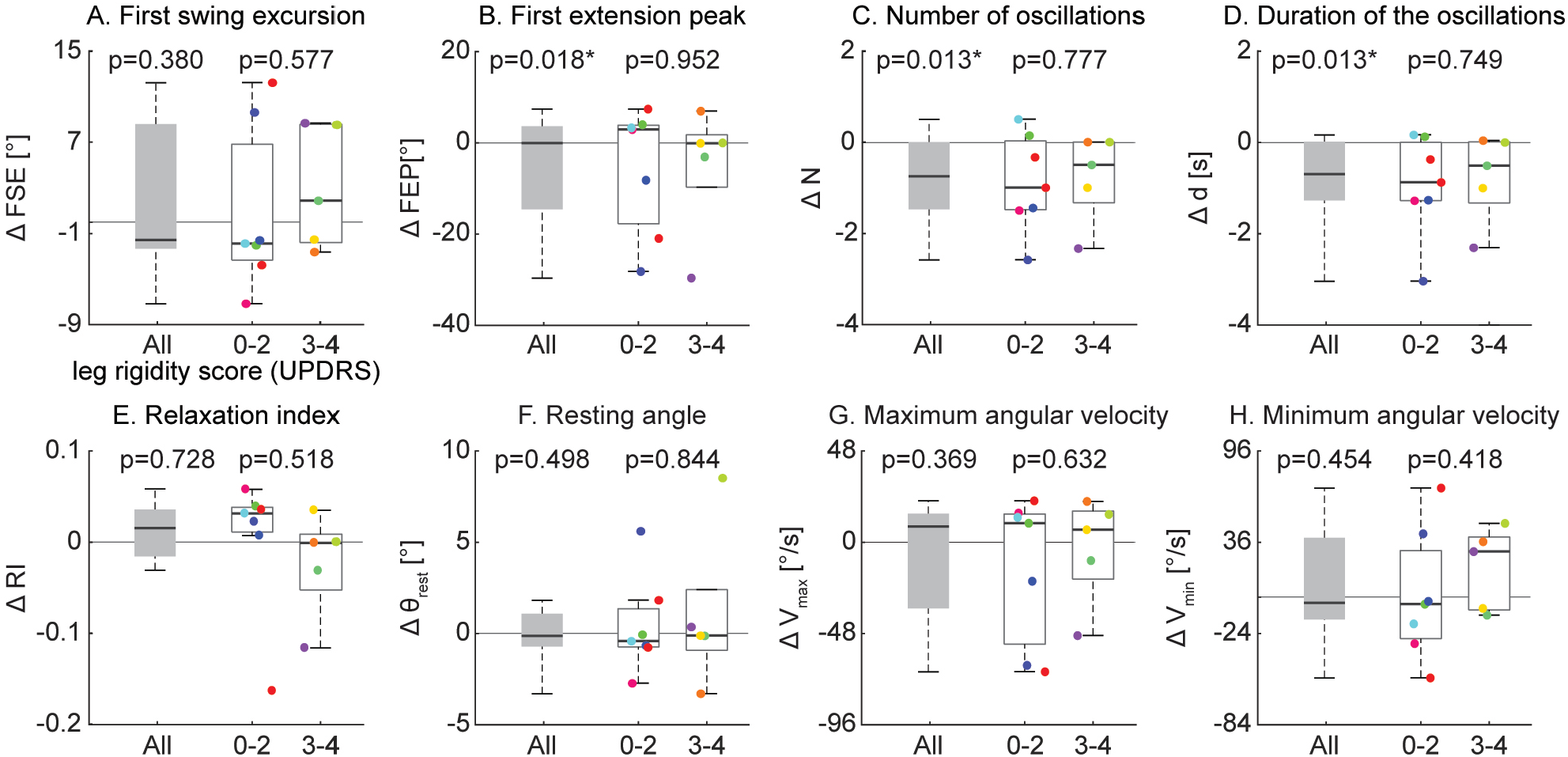
Variation of kinematic outcomes of the pendulum test during an activation maneuver. Subjects were grouped based on rigidity score of the recorded leg: subjects with leg rigidity score from 0 to 2 (absent to moderate rigidity) and subjects with leg rigidity score from 3 to 4 (marked to severe rigidity). Asterisks denote significant values (p< 0.05).

### 3.4 Fallers vs non fallers

In contrast, the effect of an activation maneuver on the biomechanical outcomes of the pendulum test were significantly different in non-fallers versus fallers (Fig. 7). In fallers compared to non-fallers, two-sample t-test revealed a significant decrease of first swing excursion (Fig. 7A, p = 0.002), first extension peak (Fig. 7B, p = 0.026), resting angle (Fig. 7F, p = 0.019) and minimum angular velocity (Fig. 7H, p = 0.026).

**Figure 7.**
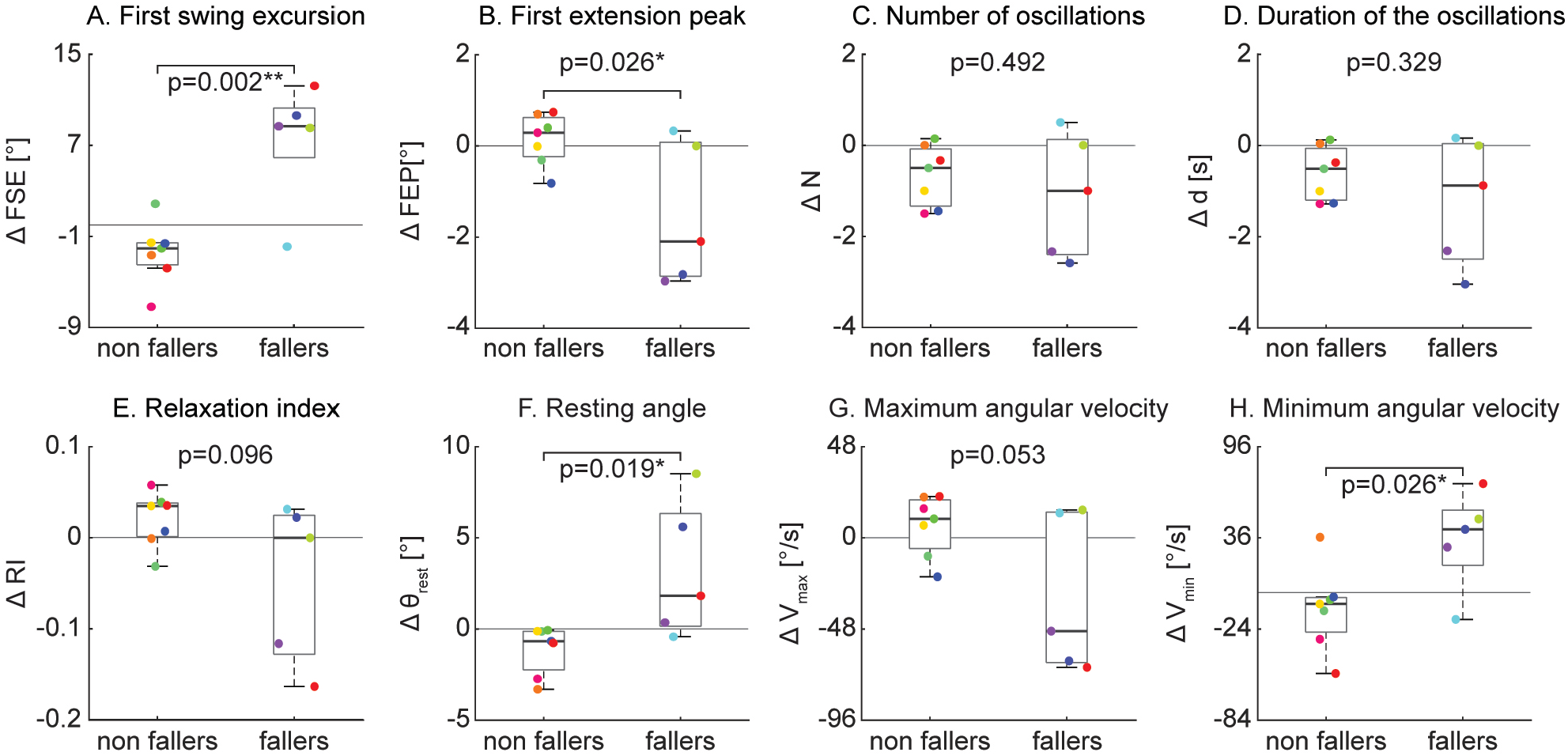
Variation of kinematic outcomes of the pendulum test during an activation maneuver in non-fallers and fallers. Asterisks denote significant values (p< 0.05).

## 4 Discussion

Our results demonstrate that the pendulum test is a valid objective measure to assess both resting and activated lower leg rigidity in people with PD. Four biomechanical metrics describing the oscillating pattern of the leg during the pendulum test were lower in those with higher leg rigidity scores, suggesting that a simple kinematic analysis of the pendulum test is sufficient to assess leg rigidity in PD. Further, in the presence of an activation maneuver the pendulum test biomechanical outcomes were altered to a different extent among participants demonstrating a sensitivity of the pendulum test to changes in rigidity. However, the effects of the activation maneuver on biomechanical outcomes was independent from the severity of leg rigidity scores at rest. On the contrary, individuals exhibiting an effect of the activation maneuver on biomechanical outcomes experience more falls in the preceding 6 months, suggesting that increased activated rigidity could be related to increased risk of falls and highlighting the need to clinically evaluate activated rigidity independently from resting rigidity. Individual differences in the changes in biomechanics and muscle activity when performing the activation maneuver also suggest that there may be diverse underlying neural mechanisms at play that warrant further investigation.We conclude that activated rigidity may play an important yet unexplored role on balance function in people with PD. The pendulum test may provide an important objective evaluation of resting and activated rigidity that may contribute to a better understanding of fundamental mechanisms underlying motor symptoms in PD and their fluctuations, evaluate the efficacy of treatments, and potentially reduce the risk of falls.

This is the first study to demonstrate that biomechanical outcomes of the pendulum test may be useful in objectively assessing the severity of leg rigidity among PD participants. A few studies have described the abnormal pattern of the pendulum test in people with leg rigidity (Schwab, 1963; Brown et al., 1988; Le Cavorzin et al., 2003), but its relationship to the severity of rigidity has not been assessed previously. Here we found that the first extension peak, number of oscillations, relaxation index and maximum angular velocity were significantly decreased in PD people with marked rigidity compared to PD people with moderate rigidity. Further, we observed that our less rigid group had altered pendulum test kinematics with respect to outcomes reported previously in healthy adults (Stillman and McMeeken, 1995), although some of the differences could be attributable to aging and require further exploration. In this pilot study, we focused on the association between biomechanical outcomes and rigidity severity, but larger studies will be required to assess the sensitivity, reliability, and repeatability in order to validate these measures for clinical assessment of rigidity and account for potential confounding factors (McKay et al., 2018).

The pendulum test has the potential to be an objective, simple, fast, practical, and affordable diagnostic method to evaluate rigidity. Expert neurologists can commit an error of up to 20% in assessing rigidity (Rizzo et al., 2016). Other instrumented clinical tests allow the evaluation of objective continuous parameters overcoming the limitations of the UPDRS rating scale, which include surface electromyography (Eisen, 1987; Andreeva and Khutorskaya, 1996), myometry (Marusiak et al., 2010), and/or torque measuring devices (Kirollos et al., 1996; Patrick et al., 2001; Endo et al., 2009; Xia et al., 2011; Powell et al., 2012; Zetterberg et al., 2015). However, to the best of our knowledge, all the methods previously proposed in literature focused on the objective quantification of upper limbs rigidity (Ferreira-Sánchez et al., 2020). The biomechanical outcomes of the pendulum test can be easily evaluated through simple observation of the leg swing or by using affordable devices equipped with gyroscope (Yeh et al., 2016) or simple video source (i.e. markerless motion capture, Mathis et al., 2018), making it feasible for standard clinical practice and telemedicine. For example, automated analysis of the pendulum test could be implemented into smartphones (Prince et al., 2018) whereas prior methods require expensive additional devices, data processing and technical assistance (Ferreira-Sánchez et al., 2020).

This study supports the idea that resting and activated rigidity should be regarded as independent variables and scored separately (Fung et al., 2000). Currently, activation maneuvers are used in clinical evaluation only to detect rigidity at an early stage, or to bring rigidity into evidence if it does not manifest at rest. In this case, the UPDRS rating system assigns a score of 1, that is not dependent on the amount of rigidity elicited by the activation maneuver, and activated rigidity is not assessed if the resting rigidity is scored at a 1 or higher. Despite several studies quantifying the effect of an activation maneuver on rigidity (Fung et al., 2000; Hong et al., 2007; Powell et al., 2011), it is not clear whether the activated rigidity is greater in people with higher resting rigidity. Here, biomechanical outcomes revealed no differences in the effect of activation maneuvers between groups with clinically assessed moderate and marked rigidity, suggesting that the effect of the activation maneuvers may be independent of rigidity severity at rest. Heterogeneity in the manifestation of activated rigidity may further provide insight into the varied mechanisms of motor impairment in people with PD. Several factors have been suggested to contribute to rigidity including an increase in involuntary background activation, changes in non-neural muscle tissue properties, increased stretch reflexes and presence of shortening reaction (Berardelli et al., 1983; Lee et al., 2002; Xia et al., 2016; van den Noort et al., 2017). Furthermore, asymmetrical patterns of rigidity can be present among extensors and flexors (Meara and Cody, 1993; Xia et al., 2009). Although we recorded EMG activity only in a subsample of participants, our exploratory results suggest that increased tonic and reflex activity could be not mutually exclusive manifestations of rigidity. Indeed, while some individuals showed an increase of tonic activity in either flexors or extensors during an activation maneuver, others had an increase of reflexive activity.

Clinical assessment of activated rigidity - even when rigidity at rest is present - could help identify individuals with higher risk of falls. The recently identified relationship between leg rigidity and falls in people with PD (McKay et al., 2019) highlights the need for more objective and continuous measures of leg rigidity (Ward et al., 1983). Here, we showed that the effects of the activation maneuver on pendulum test kinematics are greater in fallers compared to non-fallers, suggesting a causal role of activated rigidity in postural instability. Activated rigidity likely reflects a more realistic scenario of daily life, in which different concurrent tasks (such as talking or carrying an object) are performed during balance control. We found no significant difference among the tested activation maneuvers, supporting previous findings about the non-specificity of activation procedures (Hong et al., 2007). Moreover, several studies have shown that therapeutic treatments can have a differential efficacy in reducing resting and activated rigidity (Webster and Mortimer, 1977; Caligiuri and Galasko, 1992; Kirollos et al.,1996; Krack et al., 2003; Shapiro et al., 2007). As such, the monitoring of activated rigidity could help predict the functional motor impairments arising during daily activities that may lead to falls, although this relationship is still unknown. The efficacy of treatments and rehabilitative interventions aimed at reducing rigidity should take into account individual responsiveness to both resting and activated rigidity. The pendulum test could help identify the extent by which multiple impaired physiological mechanisms manifest from patient to patient, representing a potential approach to understand the functional implications of resting and activated rigidity on movement.

## Data Availability

The data that support the findings of this study are available from the corresponding author.

